# Performance management and development system in South Africa, a necessary evil. Qualitative study

**DOI:** 10.1101/2025.01.08.25320233

**Authors:** Ozo J Ibeziako

**Affiliations:** *Department of Family Medicine, Faculty of Health Sciences, University of Pretoria, Pretoria, Gauteng, South Africa; Specialist Family Physician, Head of Unit, Region 1 NSDR Ekurhuleni Health District, Gauteng, South Africa

**Keywords:** staff, performance management, health, healthcare workers, transformation and scale up, District Health System, and Primary Health Care, South Africa

## Abstract

Performance management, primarily developmental-oriented, improves individual and organisational performance as key enablers of improved services. Accomplishing performance management’s objectives are critical in the current global challenges of the shortage of healthcare workers and providing equitable, quality healthcare for all. The weakened South African district-based primary healthcare system is strongly linked to poor leadership and governance. This study aims to understand how doctors in primary health care experience the performance management and development system. The objectives explored what medical officers know about it and their experiences. Emerging themes could provide insights into improving its implementation. The study used a qualitative, interpretive phenomenological research design. Data was collected through Focus Group Discussions with semi-structured interview guide. Stratified purposive sampling was used, and thematic analysis was performed. The overarching theme was Performance Management and Development System, a necessary evil with benefits and challenges. The subthemes included understanding its multiple components, understanding clinic systems to improve service outcomes, promoting employee-supervisor interactions and interconnectedness, creating a performance and learning culture, and personal and professional growth. Effective and efficient implementation of the Performance Management and Development System at the coalface demands that managers and supervisors drive this process in an informed and strategic manner. Managers and supervisors entrusted with clinical governance should invest time in personal development to understand the process and creatively consider a fit-for-purpose implementation tool.

## Introduction

Performance Management (PM) is how work is done and organised^.1^ The Performance Management and Development System (PMDS) framework enshrines activities that influence continuous and progressive quality services through personnel and organisational reflection, correction, development, and inter-connectedness. Leadership and governance contribute to the health system’s (HS) framework and building blocks to strengthen performance and achieve health outcomes.^2^ PM is a means to an end if organisational performance links to individual staff performance.^3^ PM has become an approach to an organisation’s shared vision of purpose and aims. This is because it measures and monitors overall performance and mutually benefits employees and employers by driving staff job satisfaction and commitment to organisational goals.^3^ PMDS, if well implemented, creates an environment of "high performance, trust, and mutual understanding by defining roles, responsibilities, and accountabilities, communicating clear expectations and standards within required competencies and the expected behaviours, and establishing achievable key result areas and performance indicators.”^4^

Within the South African HS, Primary Health Care (PHC), a vehicle to deliver comprehensive, equitable, quality, integrated, patient-centred and area-based care, relies on a solid base of the District Health System (DHS).^5^ Despite the gains of PHC, such as access to health and equity, PHC performance in SA could be better. This has been attributed to structural deficiencies (weak DHS) and poor regulatory framework (lack of accountability).^6^ Leadership and governance are among the World Health Organisation’s (WHO) six health system building blocks,^6^ and could address the weakened DHS. The PMDS is a tool for leadership and governance which helps improve and maintain high standards.^7^ In South Africa (SA), Family physicians, “a doctor working as a medical generalist in the DHS and registered as a specialist in family medicine”^8^ have been entrusted with ensuring and maintaining clinical governance to strengthen DHS.^5^

According to the Department of Public Service and Administration (DPSA) policy of 2007, PMDS has determined objectives such as ensuring employees know and understand their expected job outcomes, establishing a performance and learning culture in public service, improving service delivery, promoting employee-supervisor interactions, and identifying, managing, and promoting growth in staff by addressing each individual’s developmental needs.^9^ The South African PMDS framework hinges on two dimensions: first, the Key Result Areas (KRAs) describe the broad area of staff roles and responsibilities, that is, the work that must be performed. Particular attention is given to measurable outputs - tasks, actions, and activities - that will assist the unit in effectively contributing to achieving the departmental strategic goals and objectives. The KRAs, thus, will reflect the main areas of focus of the employee from the different departmental programmes. Indicators measure the employee’s achievement of the expected outputs. Further, “the KRAs should preferably not exceed five or six substantive outputs”.^1^ Second, the Generic Assessment Factors (GAFs) are a list of generic management competencies that consider knowledge, skills, and attributes relevant to the employee’s work. Resource requirements, personnel number, job title, post grade, and the main objectives of the employees’ job complete this framework. The KRAs are weighted in percentage according to their importance level and impact on service delivery, with a minimum weight of 10% and a maximum of 30%, respectively, and should sum up to 100%. The GAFs relevant to the staff’s work area are integrated and assessed with the KRAs.^1,10,11^

Since 2018, PMDS for all government employees has been done at the mid - and end of the financial year. Assessment is scored using a four-point Likert scale against a previously agreed upon standard, performance agreement (PA) or contract. A score of one is not effective (expected standards have not been met), two is partially effective (some expected standards have been met), three is fully effective (expected standards have been met in all areas of the job), and four is highly effective (performance far exceeds expected standard). Participating in the annual PMDS ensures pay progression, while a score of four attracts cash rewards. Further, employees who score a four will submit a supporting portfolio of evidence with motivation. Appointed members of the moderation committee ascertain that the performance assessment has been done in a realistic, consistent, and fair manner that ensures the same standard is applied across the department.^9,10^

Family Medicine Ekurhuleni health district, in keeping with the DPSA directives, develops its annual departmental-specific PA or contract for doctors (Appendix I), informed by the National Development Plan (NDP) and mid-term strategic goals (MTSG) and other relevant strategic documents.^4,10^ Family Medicine Department Northern Ekurhuleni Subdistrict 1 (NSDR1), in addition, translates the expected outputs contained in the contract for doctors into a monthly practice self-assessed performance and management tool (MPST) (Appendix II) to enable each medical officer (MO) to plan, provide service, and manage practice (deployed clinic) and processes. The MPST consists of output indicators measured by auditing patient clinical records, analysing program outcomes and educational/clinical governance activities, identifying gaps, reporting successes and challenges, and creating an action plan for the next quarter. Performance is discussed individually, and best practices are shared at the unit meetings. This makes the doctor an active collaborator and driver of their performance, assessment, and areas of improvement and development.

### Problem statement

Despite over four decades of implementation of PMDS in SA^1^, it is “one of the most contested systems in the South African Public service”.^9,12^ The South African literature around PMDS has belaboured challenges around its acceptance, unfairness, unrealistic, not uniformly applied, weak accountability, favouritism, and overemphasis on its financial incentives undermine the developmental goals.^12,13^

Doctors fall within the category of middle management services and participate in PMDS. In the context of the negative discourse around PMDS and the need to strengthen DHS, one could question if the MPST influences how the doctors based at PHC experience PMDS and thus contribute positively to the DHS. This study aims to explore how MOs deployed at NSDR1 experience the PMDS. No similar research has been done in the researcher’s context. The objectives explored how MOs - “a doctor employed as a medical generalist in the DHS and not registered as a specialist in family medicine”^8^ - deployed to NSDR1 understand PMDS and their experiences. This study could shed light on the extent of the negative discourse/challenges in this group of professionals practising in a PHC setting. The emerging themes and insights could crystalise its benefits and argue the place for effective PMDS implementation strategies at the coalface.

## Materials and Methods

### Study design

A qualitative interpretive phenomenological research design was appropriate for this study because it is all about searching for meaning in how participants experience the PMDS and accurately presenting their experiences.^14,15^ Participants, MOs who experienced the phenomenon under study (PMDS assessments), were the units of analysis. The researcher not only described but interpreted the meaning of the phenomenon.^14,15^ The researcher was the primary data collection and analysis instrument, employing an inductive strategy to build meanings from the participants’ direct experiences.^14,15,16^

### Research Setting

The research was done in NSDR1, which spans the geographic regions of Tembisa, Kempton Park, and Northern Boksburg, Gauteng, South Africa. Twenty government clinics - two community health centres (CHCs) and 18 PHCs - are supervised by two Specialist Family Physicians (SFP) based at each CHC. Each clinic has at least one MO deployed to work there, and the clinic is considered his/her practice. Where more than one MO is deployed, such as the CHC, there is joint practice management. Comprehensive PHC services are provided in all clinics.

Primary healthcare facilities, comprising of PHC clinics and CHCs, serve as gatekeepers to higher levels of care (hospitals) and, therefore, are the first point of contact between the community and healthcare.^17^ Currently, the PHC and CHCs are organised and function guided by the Ideal Clinic Framework (ICF) launched in 2014 through Operation Phakisa. The ICF has a double aim: address deficiencies in primary-level care and systematically transform it to conform to the National Health Insurance (NHI) standards. An ideal clinic is a “clinic with good infrastructure, adequate staff, adequate medicine and supplies, good administrative processes and adequate bulk supplies that use applicable clinical policies, protocols, guidelines as well as partner and stakeholder support, to ensure the provision of quality health services to the community.”^17^ The ICF proposed a comprehensive package of healthcare services from pre-birth to death, health promotion to palliative care, and a level of care from the community to the district hospital. Integrated Clinical Services Management (ICSM) became the key focus within the ideal clinic. ICSM is organising the clinics so that “people get the care they need, when they need it, in ways that are user friendly, achieve the desired results and provide value for money.”^17^ ICSM should be a seamless integration of services. All services are offered daily and organised in four streams - chronic, acute or walk-ins/emergency, mother/child and sexual reproductive health, and therapeutic services - according to set standards to achieve patient satisfaction. All streams except acute/emergency services are planned with scheduled appointments. Monitoring and evaluation to maintain ideal clinic standards use a dashboard of components (10), sub-components (32) and indicators (273).^17^

A CHC, like the research setting, differs from a PHC because it provides 24-hour services, including a midwife-obstetric unit and additional services such as a secondary psychiatric clinic run by the psychiatrist and MOs in the team, dental clinic and radiology services. The workforce at the CHC and PHC is multidisciplinary, consisting of doctors, nurses, and assistant pharmacists. Doctors and nurses dispense medications from the consulting rooms because it is outside the scope of practice of the assistant pharmacist. The therapeutic services professionals include physiotherapists, occupational and speech therapists, audiologists, podiatrists, dietitians, clinical psychologists, and social workers outreach on different days of the week to the CHC and PHC.

Further, this region has only one hospital, which, although upgraded to a tertiary level, still functions as a district, provincial, and tertiary hospital. Doctors employed at the CHC and PHC are of different grades of MOs (Grades 1 to III), community service medical officers (CMOs), and medical interns rotating bimonthly during the two months of Family Medicine rotation in the PHC and CHC. The MO consults all patients referred by nurses from any of the streams in the clinic and refers difficult cases to the SFP / hospital when necessary. In addition, stable patients are booked for chronic care and antenatal visits by the doctor. The expected role of the MO mimics those of the SFP - a clinician; clinical trainer of other junior doctors, nurses and community health workers; some level of clinical governance; and a team player in community-oriented primary care (COPC).^8^

### Population and sampling

Seven community service and 14 full-time MOs were the staff complement of doctors in NSDR1 during the study period. Eighteen out of 21 MOs working at NSDR1 who met the selection criteria participated in the study. The inclusion criteria were at least one year of employment in the unit and participation in the mid-year and annual PMDS of 2022-23. One year is the minimum time frame to participate in the annual PMDS, which includes submitting a portfolio of evidence if the employee scores four.

Stratified purposeful sampling was used, and selected participants were grouped into four strata according to the duration of their employment in the unit: one year, one to two years, two to three years, and above three years. Participants were contacted telephonically. Four MOs did not fulfil the selection criteria because they had been employed for less than one year and had only participated in the mid-year PMDS assessment. Ethics approval and permission to collect data were obtained before data collection.

Information leaflets and consent forms were forwarded to all MOs (18) who met the selection criteria and gave verbal consent. Participants signed a written consent after perusing the information leaflet and before data collection. Focus Group Discussions (FGD) were held between October to December 2023.

### Data collection

A semi-structured interview guide (Appendix III) was appropriate for exploring participants’ lived experiences. The guide was piloted and validated by conducting an individual interview. No changes were made in the wording after the piloting was completed, confirming its accuracy and clarity. Results from the individual interview have not been included to ensure anonymity.

Four FGDs of two to six participants were conducted. The number of participants in the FGD was determined by the number of years of employment in NSDR1, the criteria for inclusion in the strata. The purpose of stratification was to elicit divergences in how this phenomenon was lived. Using a semi-structured interview guide, the researcher conducted the FGDs at the participants’ convenient time and comfortable venue. The aim of the research was explained once again. The researcher encouraged participants to express themselves freely and sincerely as each lived the experience. Participants were cognisant of the importance of respecting each other. The questions were not asked sequentially but were determined by issues raised from the preceding discussion point. The researcher is one of the SFPs in the unit. A research assistant (RA) was trained in the event of a power dynamic, which did not arise. Data was collected until saturation was attained with the last group discussion.^15,18^

### Data analysis

The researcher transcribed the FGDs verbatim using the Descript AI software. Subsequently, the researcher listened to the audio repeatedly while correcting and completing the transcripts to ensure authenticity. Thematic analysis was done using the ATLAS.ti software. Thematic analysis was preceded by repeatedly listening to the audio, re-reading and reviewing the transcript for corrections and meaning, described as “getting the sense of the whole”.^14^ Subsequently, the transcripts were imported into the ATLAS.ti software where recurring words, phrases or sentences describing participants’ experiences were initially coded as the first step in thematic analysis.^14,18,19^ This has been described as the “meaning unit”.^14^ Meaning units or generated codes were categorised as the second step,^14^ then sub-themes, and finally, themes emerged as the last step of thematic analysis.^14,18,19^ The researcher is familiar with thematic analysis from previous research experience. In addition, codes and themes generated were contrasted with peer review.

### Ethical considerations

Permission for this study was granted by the National Health Research Database (GP_202309_001) and Ekurhuleni Ethics Committee (10/07/2023/01). The University of Pretoria Health Sciences ethics committee granted ethical clearance to conduct this study (No. 458/2023).

### Trustworthiness^19–23^

Trustworthiness refers to qualitative research’s "quality, authenticity, and truthfulness."^20^ Lincoln and Guba described credibility, dependability, conformability and transferability as the four pillars of trustworthiness. *Credibility* means the congruence of the findings with participants’ lived experiences. The researcher achieved this through triangulation, using multiple information sources such as repeated reading of transcripts and member checking of data and its interpretation with participants to establish identifiable patterns. *Transferability* concerns how applicable the findings are to a new context. This was achieved in this study by providing a detailed description of participants and the research process (study context, sampling, demographics, selection criteria, interview guide, and analytical steps), and data collection continued until saturation. By the end of the fourth FGD, data saturation was deemed attained because FGDs, transcriptions and analysis ran concurrently, and no new information or theme emerged. *Dependability,* which looks at consistency, was achieved by peer debriefing and keeping the researcher’s experience and interpretations (bracketing) at bay. *Conformability* means neutrality to ensure that findings are grounded in the data. This was ensured in this study by keeping a tight audit trail for data collection, analysis, and interpretation.

### Reflexibility^20,22,23^

The researcher, one of the SFPs employed at NSDR1, has experience in qualitative research. The researcher has no interest in PMDS and participates in and implements it. Given the human resources department’s tenacious effort to ensure PMDS is completed annually, the researcher has self-interrogated its benefits, challenges, and shortfalls. The researcher actively engaged in critical self-reflection on potential biases and pre-conceived ideas that might influence data collection, analysis and interpretation. The researcher did not share feelings or perceptions during data collection because the researcher did not in any way express or insinuate the personal lived experiences regarding PMDS. Also, the researcher focused on participants’ lived experiences during analysis and interpretation by drawing out meanings expressed by participants, and therefore, was non-judgemental and non-directive. There was a research assistant should power dynamics arise. Participants showed honesty, transparency, and openness while sharing their feelings, perceptions, and experiences. The research results were provided to participants for scrutiny, corrections and amendment as deemed necessary.

## Results

### Participants Characteristics

The demographic non-personalised information of age, gender, duration of employment with the Department of Health (DoH) and Ekurhuleni Health District, and the number of years participating in PMDS cycles were collected from participants. Seventeen MOs were interviewed - (community service, four [23,5%]; full-time, 13 [72.22%); and four (23,5%) were males, while females were 13 (72.22%) with a mean age of 41 years. In their first employment as independent practitioners, CMOs, comprising most FGD1 participants, were the least exposed to the PMDS process (4 [23,5%]). PMDS experience was not parallel to most participants’ years of employment with the DoH. Table 1 shows participants’ de-identified demographic characteristics.

**Table 1.** Table on the Participants’ De-identified Characteristics.

| **GROUP** | **MEAN**<br>**AGE** | **MEAN**<br>**EMPLOYMENT**<br>**DURATION**<br>**(\*GDoH)**<br>**(YEARS)** | **MEAN**<br>**EMPLOYMENT**<br>**DURATION**<br>**(\*\*NSDR1)**<br>**(YEARS)** | **MEAN**<br>**PMDS**<br>**CYCLES IN**<br>**(\*GDoH)**<br>**(YEARS)** |
| --- | --- | --- | --- | --- |
| FGD1<br>(6 participants) | 30 | 3.6 | 1 | 1 |
| FGD2<br>(5 participants) | 59.2 | 16.6 | 14.2 | 6 |
| FGD3<br>(2 participants) | 31 | 5.5 | 2 | 2 |
| FGD4<br>(4 participants) | 40.25 | 7.75 | 3 | 3 |

\*GDoH, Gauteng Department of Health
\*\*NSDR1, Family Medicine Department Northern Subdistrict 1 Ekurhuleni Health

### Themes and Sub-themes

**Fig 1.**
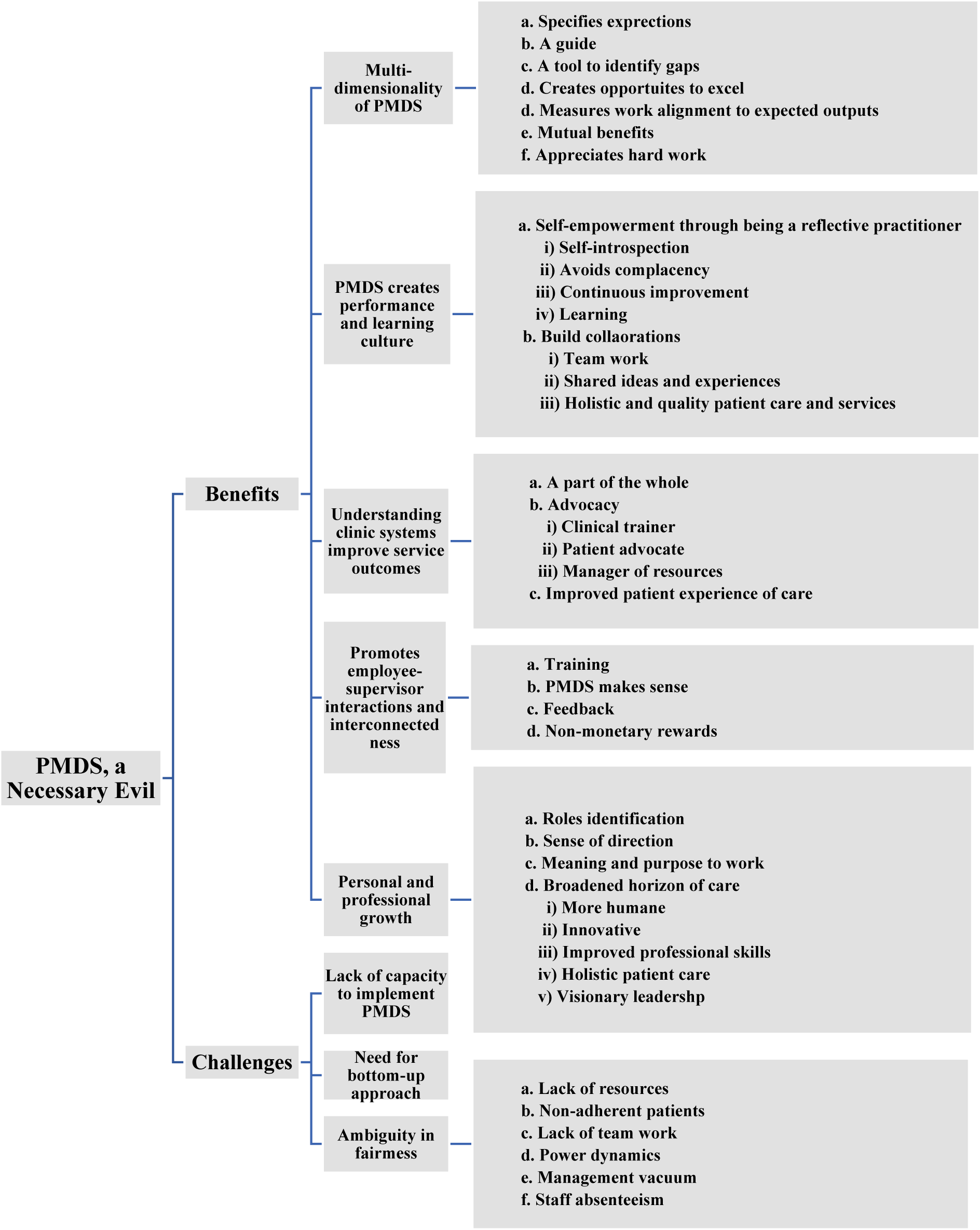
Themes and Sub-themes.

#### 1. PMDS, a necessary evil

Overall, participants expressed depth in their knowledge and understanding of the PMDS. PMDS, “a necessary evil,” was the insightful overarching theme articulated by FGD2 of PMDS-experienced participants. Generally, participants elucidated PMDS’s multiple components, experiential benefits, and undeniable challenges.

##### 1.1 Benefits

###### a. Specifies expectations, guides, a tool to identify gaps, and create opportunities to excel

The participants from all FGDs highlighted that the contract within PMDS is a tool that guides, assesses, and measures the employee’s compliance or alignment with expected outputs. Although arduous, progressive learning by doing clarified PMDS’ necessity and usefulness because the contract outlines what is expected of the doctor, especially for doctors new to primary health care and young doctors. Participants highlighted the importance of guiding a doctor’s job expectations in the primary care setting, which tends to present as a novel work environment because medical training is mostly hospicentric.

> ***FGD2****: “Maybe for us it’s obvious, our roles, because we have been for a long time in this business. But for the new doctors, the person that comes first to work in the clinic, to work in primary care, they won’t know if they don’t have something to guide them on what to do.”*

The PMDS, as a tool and guide, has output activities regarding what patient care entails in practice. It allows the doctor to strive for and attain the expected clinical targets and standards of care.

> ***FGD1:*** *“It helped me to, in terms of how I should see my patients. I know I need to do my annual eye exam, foot exam, that’s how they should be seen; that’s the standard that you are setting. It gives you a sort of a guideline. I think it helped me in that sense to have a PMDS and not feel lost in the system as well.”*

###### b. Measures work alignment to expected outputs and appreciates hard work

PMDS assesses and *measures employee alignment with expected outputs*, *appreciates hard work*, and improves the quality of patient care. The MPST included output indicators with set targets for each quarter. These targets guide the doctor in gauging what needs to be done, how, and when. It provided an efficient and effective monitoring, evaluation, and implementation tool at the coalface. The result was *mutual benefits* for patients and employees.

> ***FGD4:*** *“…It’s lovely tool (MPST). it keeps us in line. It encourages us…to do better; to do more; be reviewing and be checking what we’ve done, if it has benefited us in the way that we wanted to benefit. Again, if there are gaps…, keeping us abreast with our gaps. Where were your challenges? What have you done to try and mitigate those challenges? It’s a tool that is guiding…where we are going in terms of how we are performing, trying to close all the other gaps that we sometimes encounter in our workplaces.”*

All participants clearly expressed the linearity between meeting expected outcomes and cash reward according to the DPSA policy, which is a good reason to ensure that contract activities align with the expected outcome. For participants, this meant working in a directed and effective way and sometimes working under pressure to get things right. Some participants also expressed that it is an assurance that the work was not done in vain.

> ***FGD3:*** *“…linking it up with the contract, we have certain specifications on what we are expected to do. The PMDS is just sort of a measure to make sure that we are in line with that so that we are able to identify it (gaps) or not, then we can find ways to sort of improve on that. And if you’re performing, …get compensated for the performance.”*

> ***FGD4:*** *“…my understanding with regard to PMDS is that it is a tool that is used by the employer to try and assess performance of all the employees. But before that is done, there’s always a baseline that is given in terms of the duties that one has to fulfil. Based on that, you check yourself more, you weigh yourself according to what is put for your baseline and what you have actually achieved at the end of the day. So just to encourage people to perform, that’s number one. But also is a means to ensure that you appreciate those that have performed beyond what is just given. It’s more like an encouragement because at the end of the day, it’s got some monetary, …awards or rewards that will actually motivate employees to always go above and beyond what they’re supposed to give in order to achieve the goals?”*

###### a. Self-empowerment through reflections

For the junior doctors who are PMDS inexperienced, the contract activities and expected output meant a lot of work. They were exposed to clinical practice differently. Becoming a *reflective practitioner* involves *self-introspection*, *avoiding complacencies,* taking the easy way out, and *being aware of the need to improve and learn continuously*. Reflecting was critical to self-empowerment.

> ***FGD1****: “…It has helped me understand the way I work and revealed areas that I needed to improve on, like being more organised and consistent. I’ve also had to work under pressure and put in extra effort, take work home, interpret and make sense of a lot of new information; that was a stretch for me but I tried my best to get things right so I guess I am open to continuous learning and new challenges. …The assessment exposed me to a lot of things, so I’ve had to step up, empower myself through learning and adjust quickly. Work became less stressful, and I’ve become more comfortable and efficient.”*

> ***FGD2:*** *“In terms of self-introspection, how do you do a holistic approach to your patients?…”*

> ***FGD3:*** *“The one is that I think it stops people from being complacent. Because I think when you don’t assess people, then people can just do, like not even the daily amount. They’ll just do whatever …as long as they get paid. So, I think it picks up on that.”*

Participating in the PMDS meant examining what and how they do their work and striving to give their best. The result is a continued willingness to learn, get out of one’s comfort zone, and improve oneself and one’s work.

> ***FGD4****: “…I would say, at first, I was petrified because I come from… setting, and it was only one department I was assisting. Now it was like all the departments.…When I arrived, the very first time, it was like more serious you know…As time goes on,…I get to know that I learn from it, and I got to know that I have duties, I have to follow the job description…To know what’s happening in your clinic…Because if it was not for the PMDS,…I’ll probably not be involved with certain things like knowing what’s happening in data room, and the statistics and how they do things.”*

###### b. Build collaborations through relationships to improve patient care and services

Learning is enhanced in a collaborative environment. All participants emphasised the importance of teamwork and building relationships, expressed as a bond. The synergy between doctors and clinic teams enhanced intra-facility communication and referrals.

> ***FGD4:”.*** *Like,… Phuthuma, viral loads, and they know that I’m also interested in knowing, and….they must be on their toes. I need to know what’s going on. If a patient is not suppressed, they must immediately refer to me and also those tracing the patients…So they know that they have to be on their toes. And also the elderly patients…being seen by WBOT, they know that they must report to me; it’s not just them doing their thing.”*

*Holistic and quality patient care and services* can only be realised through professionals working together to improve health outcomes through *shared ideas and knowledge*. This includes working in teams and harnessing knowledge and skills from colleagues and other professionals, which provides positive performance and a learning culture. Participants admitted to understanding how each team within the clinic worked and learning new skills from them.

> ***FGD1:*** *"I also realised that it takes teamwork to complete PMDS because everyone in the clinic contributes to the statistics in one way or the other. I’ve had to work with different teams in the clinic like the WBOT, ANC and MOU team, TB team… And with each team understanding how they work e.g TIER.NET; HAST; how we are doing as a clinic; what gaps are there, and ways in which we can improve e.g ESMOE drills and personally also learning new skills.”*

###### a. Being part of the whole

All participants alluded to seeing the bigger picture of the whole facility and realising that contributing to it helps appreciate one’s work. In different ways, participants expressed feeling that their medical training and work were focused on clinical stuff. However, with PMDS, they recognised that patient care works through a system that needs to be in place and functional, like ensuring the availability of medical stock in the pharmacy and, when there are stock-outs, what processes should be followed. By working with clinic teams, one is also informed about what is happening in the clinic’s other services.

> ***FGD3:*** *“…it (PMDS) helps me to see the whole clinic or where I’m working in a bigger picture; that this is going on, this is happening there. You have a better understanding of what’s going on in the whole facility…. It helps you to understand the institution as a whole, and then it gets much easy for you to work when you know that, when I need this, I need to go there… How is the service going…”*

For the participants, being part of a whole also meant experiencing firsthand how clinical activities at the coalface contribute to global health data, which inspires and appreciates one’s work.

> ***FGD1:*** *“…It helped me realise that I can contribute to the statistics and that all we do contributes to us as a country, meeting targets set out in the sustainable development goals. Realising this contribution made me more appreciative of my work and its contribution.”*

###### b. Advocacy

Advocacy becomes a consequence of acknowledging one’s role in a system. PMDS-experienced participants recognise that their roles as *clinical trainer*s of junior doctors, nurses, and other healthcare professionals, *patient advocates*, and *resource managers* impact the health system and the quality of services. Educating other team members empowers them and creates a joint vision in clinical care using limited resources.

> ***FGD2****: …”And your role as well, to be the advocate of government’s resources.”*

> ***FGD2:*** *“I now make Wednesday topics that is about medicines that usually we prescribe. I start with Lanzoloc. I already did prednisone. Because I have one or two sisters who order prednisone for flu or for arthritis… I said, every Wednesday we’re gonna talk about different medications that are wrongly prescribed.”*

Exercising a patient advocacy role meant the doctor ensured patients accessed the appropriate level of care and awakened responsibility and accountability among other clinicians and colleagues.

> ***FGD2****: “…I sent a patient with COPD, a very severe COPD to General OPD. This patient was just seen and given Asthavent and was not given even any date. I phoned…, I said, ‘ah, this is unacceptable.’ The sister said, ‘no, no, the doctor who saw this patient is usually very thorough’…I said,… ‘I want something written down, and I want something to come back.’ Hey, he (the patient) destroyed his life and everything…But now, let’s just prolong his life a little bit, with dignity and quality of life.’ When I said that, it’s like, ‘who are you? Where are you phoning from?’…‘Okay, doctor, I will ask the doctor to see the patient’… They saw the patient again. And the patient came back. Now he comes to collect the Symbicort.*

> *He is improved, guys. He’s improved.”*

###### c. Improved patient experience of care through effective systems

Participants indicated the usefulness of clinical audits included in the MPST with varied examples, prompting the active use of internal resources, interconnectedness, and effective strategies. Good patient care, especially chronic care, is achieved and sustained through on-going monitoring of patient progress. Improved professional skills through upskilling oneself and thinking outside the box positively affect patient care experience.

> ***FGD2:*** *“…I was looking at the fact that, during COVID, a lot of patients were taken out of the system and either put on CCMDD without even seeing doctors. Those patients had never seen doctors. So you find them, like, once in a while…What the PMDS has done, it has brought into my thinking, I’ve had to think outside the box of what to do with those patients… What I then devised was a plan,…I go into WBOT and actively call…all those patients and see what I can do for them. And because of that, I’ve seen so much improvement…”*

> ***FGD1****: “… it also pointed out the shortfalls. If we didn’t do a PMDS, I wouldn’t know where my shortfalls are. For me, the referral system was heavily short-falled. It took a while, but eventually,..having to tell the sisters to call even if I am not duty on that day. Call me and discuss a patient that you want to refer. Cause sometimes you find that it doesn’t need to be referred, you can actually manage it yourself.”*

Participants articulated that improving the patient’s care experience entails paying attention to little things in clinical practice that might not require much effort but are meaningful for the patient. The biopsychosocial approach - taking time to talk to patients about their psycho-socioeconomic situations, screening and health promotion, and educating patients on lifestyle changes, empowers them to be responsible for their health; thus, patient satisfaction is assured.

> ***FGD1:*** *…"I think the greatest win for me this year was realising how merely screening for mental health, which is often the big elephant in the room, changes a lot of things. I have definitely shifted my style of consultation and have been appreciative of having witnessed its impact on a patient’s life, their attitude towards treatment, and their response to treatment.”*

Improved patient care experience was described as individualising patient care to address differing needs from a biopsychosocial approach and developing clinical care tools such as diabetic diet charts to enhance patient care. Participants became more cognizant of the social determinants of health and how they could impact patients’ adherence to treatment goals if not considered.

> ***FGD2:*** *“…if we talk about chronic patients,…the first thing that it (PMDS) has improved me is to individualise every patient… Even the ones that are doing well, I must take them as individuals. I… write SOPs. I’ve designed my own charts for my diabetics…I have got a way of talking to my patients. The approach is not always the same with every patient. Psychosocial has been my main thing that I use with my patients - the state of their poverty, the state of their wealth, the state of their families, and economics. Without PMDS, I don’t think I would have looked. I would just have gone through a patient, and as long as the next is coming. Now, I don’t look at the patient as a burden, as someone who is kind of, oh, what is he going to say today?”*

###### 1.1.4. Promoting employee-supervisor interactions and interconnectedness

Participants agreed that PMDS *makes sense* and is possible if the supervisor provides training, guidance, and tools to make it understandable and meaningful. *Training* on the MPST at the beginning of the year, especially for junior or new doctors, guided practice, and the *feedback* loop enhanced *employee-supervisor interconnectedness*.

> ***FGD1:*** *“…while I was working on my first PMDS…I was still trying to figure things out. I only realised what was expected of me when I was handing in my first PMDS assessment. There were things I needed to correct and data that I had to interpret differently. That’s when I was guided through the process and had a better understanding. The meeting we had about PMDS also cleared a lot of things."*

> ***FGD4:*** *“To me, PMDS, as a junior medical officer,…when I…was introduced to it, it was something new. So, it was a lot of work…But with more understanding and being in the system, and understanding how the system works, then it started making sense.”*

An employee wants to improve and make a difference if the goal is clear and the process makes sense. The training and follow-up meeting also provided a platform for benchmarking and learning from other colleagues.

> ***FGD1****: “I feel like PMDS does make sense as a doctor. I am not sure if being a community service doctor really entails PMDS. But definitely, after that (meetings), it makes more sense because, at the same time, you are improving yourself as well as the standards of the clinic, and at the end of the day, all we want is better patient care.”*

Participants in FGD2 and FGD4 described non-monetary rewards such as certificates of appreciation, feedback, and patient appreciation as boosting staff morale. More value was attached to professional prestige and growth, patient respect and feedback, and patient satisfaction with services received than monetary compensation.

> ***FGD2:*** *“…It’s not about the money. And I’ve always said…it’s not about the money. It’s about how I manage my patients. And the feedback that I get…I feel so good when I get feedback from my patients…”*

Participants in FGD4 also acknowledged that non-monetary forms of appreciating employees’ commitment and dedication will drive ethical and professional practice.

> ***FGD4:*** *“…I think it (PMDS) should be..put up nicely because it sometimes tends to promise what…the department cannot really do for the employees. It just should be used as a tool and not promise any type of reward other than certificates, not the monetary one. Because that one..makes people..cut corners in order to achieve your best scoring because…they want the money. Rather than…being given money, can we then get…. certificates where you’re appreciated for the work that has been done? …It will take away some of the issues that we’ve…observed…what people will do to try and get money… when they have not done justice in terms of what their job description is.”*

###### a. Roles identification, sense of direction, meaning and purpose

Personal and professional growth was welcomed as an uncomfortable aspect of growth triggered by doing the PMDS. These were driven by *role identification*, a *sense of direction*, and finding *meaning and purpose to work*. Although PMDS with the MPST put much pressure at work due to the many new things one had to learn, it changed one’s attitude toward work and made work meaningful, motivating, and satisfying. Participants allude to learning to prioritise and set work goals and identify and develop technical skills such as becoming computer literate.

> ***FGD3****: “…It helped me identify that I…have a bigger role than I initially thought. Because with the previous place where I worked, I used to just…be a clinician on the ground, do my work, see patients, and go…As soon as we got here and did the PMDS, then we started recognising that….there are some things that needs to be done in the clinic and that progress requires us (doctors) to…do them and plan..and move them forward..Like..planning on drills,..educating other team members, that rely on us…We saw that we are the ones who actually can do it. …Things like implementing BMI, which we didn’t have before, it was a matter of..talking to the staff members..to say that the equipment is there, so let us get working so we can..start implementing and doing better on PMDS.”*

> ***FGD4:*** *…”yes, it has helped in terms of putting priorities at work…according to how your performance was on the previous assessment in order to cover for the gaps…”*

> ***FGD4****“PMDS actually sets goals for you…. it helps you plan your daily work on a daily basis and then achieve what you need to achieve.”*

> ***FGD4****“…it has helped me, where you are limited,… and then what you need to really improve on things like…, being computer literate…”*

Participants in each FGD explained that identifying one’s bigger role as a doctor and planning, supervising, and implementing changes with team effort becomes a game changer in approach and reason to work. Although daunting to a young doctor, with support, it drives creativity and growth.

> ***FGD4:*** *“This one (PMDS) is quite new. How do I even meet people who are senior to me and give feedback and say, guys, here we are not doing well. So… (PMDS) also helped with being in the front line and supervising everyone, and that’s one thing that I was lacking - being a supervisor to interns, nurses,.. guiding everyone. And I’m still young…it’s one of my roles, my duties, and I really need to help all these other healthcare workers.”*

In another FGD, participants described some negative experiences of lacking meaning in work as boredom, disappointment, and feeling bad about going to work.

> ***FGD2:*** *“… the fact that if you go to the clinic - and I have had that experience from years before - if you go to the clinic to kill the queue, to finish a thing, you feel so bad going to work like that. You feel bored, you feel disappointed, and you don’t like your work, you want to do other things…”*

###### b. Broadened horizon of care

There is reciprocity between personal and professional growth because PMDS makes one *more humane* and *innovative*, *improves professional skills*, and *results in holistic patient care*. *Professional prestige and respect* draw from professional and personal growth. For the participants in FGD2, broadened care horizons include empathy, seeing the patient as a person, and seeking health information and innovative ways to engage the patient and improve care.

> ***FGD2:*** *“PMDS has improved me as a human being as well, to be empathetic,… not sympathetic…We must never be sympathetic to our patients…”*

> ***FGD2:*** *“…you find yourself looking for information, and you find a lot of things that you would never be informed because it was maybe discussed at another level…For instance, …there is a program that is ‘welcome back patients to care’. That is, never reject the patients… The patients are our team. Just welcome them back…. ‘Thank you for coming;’ ‘I’m happy that you’ve come;’ ‘You didn’t come, let’s see the reasons’. …It’s changing the approach to the patient…I learned that because I was looking for how to improve this thing (defaulters). Because we were having a lot of defaulters. So, it makes you look for information to improve your treatment, your management as well.”*

As alluded to in all FGD, one not only becomes a manager but acquires *visionary leadership* because one goes ahead of the team, foresees, plans, influences, and effects changes. Performing PMDS helped participants understand their roles and responsibilities, which extend beyond the doctor’s consultation room. It broadened the horizons of care and practice.

> ***FGD1:*** *“It has helped me see things beyond just being a clinician. I’ve had to learn about the social issues people face and understand the impact they have on patients’ health and wellbeing and correlate it with the statistics we have on different outcomes. I’ve had to learn about leadership,… different factors that contribute to service delivery and my role in all of that. I also identified things I enjoyed more at work like women reproductive health and PMTCT. Having to do different things broadened my understanding of my role as a doctor at a PHC level.”*

##### 1.2. Challenges

Participants did acknowledge that despite PMDS’s multiple benefits, there are challenges to its efficient and effective implementation that managers could mitigate.

###### 1.2.1. Lack of capacity

Managers in some units appear to lack the capacity to implement PMDS due to a lack of understanding and ability to do so. As participants in two FGDs alluded to, sometimes PMDS scores are ‘cooked’ or made up solely to get monetary rewards. The consequence could be that PMDS does not reflect employees’ actual performance but becomes mere compliance.

> ***FGD4:*** *“As a junior medical officer,… I never even thought of completing PMDS. But where I was at the time, we had an option of not completing. It means that our managers at that point in time did not understand what the purpose of PMDS was. And then from there, as I grew.. in the career,..they would complete it for us and just let us sign…It was more of a tool just to comply with what the employers wanted..We just go and sign. No proofs..They’ll just give you a **‘3’**, and you’ll be content with some satisfaction because you do not want to go beyond what you are supposed to do…I think most of the attitudes developed from that going forward.”*

Participants expressed that the managers’ lack of capacity to manage PMDS is evident in employees’ scores beyond objective performance, which are misaligned with clinic performance.

> ***FGD1:*** *“PMDS should be a true reflection of what’s happening in the clinic. There should be accountability and evidence for scores that are given, and people should have an understanding of what different scores mean. The statistics should correlate throughout the year and with any significant change, be it a drop of improvement. There should be factors identified that contributed to that, and somehow, that should make sense. What’s on paper should be what’s on the ground.”*

###### 1.2.2. Bottom-up approach

Participants in FGD1 questioned what informs the PA targets because they sometimes exceed the reality of a particular facility, such as its patient population or socioeconomic status. In some instances, a bottom-up approach to setting contextualised, realistic, and feasible PA standards involving the employees on the ground could make targets more plausible.

> ***FGD1****: “It is a good way, but performance assessment can’t be one-size-fits-all because working conditions are not the same for everyone. For some people, given the resource constraints they face or socioeconomic factors affecting the populations they serve, it may be difficult to reach the set target, which means no matter how hard they work, they may never reach the targets and may never get remunerated.”*

###### 1.2.3. Ambiguity in fairness

Participants highlighted factors out of their control, such as *lack of resources* (equipment and staff shortages) necessary for the work to be done well and *patients’ non-adherence* to clinical care targets. *Lack of team effort* because quality patient care is multidisciplinary, resulting in a low PMDS score, reflects the team not working together despite the individual employees’ efforts. *Power dynamics* between facility managers, the clinic administrative managers, and the doctors tasked with clinical governance make it challenging to bring about change to attain set standards. All these factors create a questionable situation of *ambiguity in fairness*.

> ***FGD3:*** *“.. it makes sense why it’s (PMDS) there, but I feel like for our work environment and the resources that we’ve been given, and then the targets that we are given, it’s not fair…because…we’re basically almost making a plan every day for most patients…”*

> ***FGD2:*** *“…For instance,… my clinic…because the manager retired,… we don’t have a manager…So, we don’t have good evaluations in the ideal clinic…I’ve been trying to help them and tell them, let’s do quality meetings, but it’s not in my power… I believe that doctors need to have a little bit of more power in the decisions made in the clinic. They have to consult us and ask us things… Because they do their own things, and doctors do their own thing."*

> ***FGD1****: “…some areas that are not fully reliant on you as a clinician…It’s teamwork, and sometimes you have people that no matter how much you tell them,… they still don’t do it, and… you… score lower. Then it’s not a reflection on you, it’s a reflection on the group…”*

> *Staff absenteeism and the non-replacement of retired managers create a management vacuum that increases the workload. Consequently, the clinic doctor adapts daily set work plans to compensate for the staff shortage.*

> ***FGD2:*** *“…It’s just that the situations in our clinics,… are not ideal, in the sense that it’s overwhelming, the work that we do…Like we were talking about absenteeism. On a day, sometimes, you have to intervene and help out, even beyond the number of patients that you’ve been booked for. Because, in any case, patients have to be serviced. They are here now. And there’s only one chronic sister. Such things, then you get… derailed… in whatever you want to do….”*

## Summary of Key Findings

PMDS, a necessary evil, was the overarching theme in this study. The challenges of the manager’s lack of capacity to implement the PMDS, the need for a bottom-up approach to set performance agreement output targets, and ambiguity in the fairness of who gets rewarded were indisputable in this group of professionals and primarily out of their control. However, PMDS’s outstanding benefits articulated by participants seem to support its role in helping public servants be responsible and accountable, grow as individuals and professionals, and contribute to the organisation’s goals.

## Discussion

PMDS’s multidimensional benefits in this study align with the DPSA’s^9^ goals of the PMDS. Although factors may be beyond one’s control, PMDS implementation strategies could positively impact employees’ experiences, job satisfaction, and development.

### Training with support provides knowledge and appreciation of the PMDS

Participants expressed an in-depth understanding of the PMDS. They appreciated the unique place of having a PA or contract that outlines agreed-upon activities directed towards expected outputs. Training and guided practice enabled participants to articulate the why and purpose of PMDS, in keeping with some definitions of PM by Aguinis as a continuous process of identifying, measuring, and developing the performance of individuals and teams and aligning performance with the organisation’s strategic goals.^24^

MPST guided participants on the expected output indicators. Training at the beginning of the financial year, quarterly submissions of MPST reports, and meetings to share best practices enhanced its use. The MPST enabled participants to self-assess, monitor, continuously correct, and implement changes. Studies have highlighted the critical role of training, which informs and empowers managers and employees but is often lacking.^25,26^ Training on the PMDS becomes effective if there is practice guidance. Lockwood argued that employees are more productive if they have the necessary knowledge, training and development to do their jobs.^27^ An eight-point managerial PMDS implementation strategy described by Sambo and van der Walt still leaves a vacuum in its practical translation at the coalface.^28^ The positive impact of the MPST is significant because Melnyk et al.^29^ indicated that PM could adversely impact performance when misalignment exists between what the organisation wants to achieve and what is being measured.

Participants accentuated the ongoing nature of PM, which is opposed to performance appraisal, a reductionist approach to the essence of what PMDS is supposed to achieve. Smither and London^30^ punctuated that managers who lack this understanding make PM a mere tick-box exercise. Also, for Aguinis, evaluations done twice a year without constant coaching and feedback to the employee are just performance appraisals and not management.^24^ The fact is that performance appraisal should form part of PM.^24,31^ In the study by Govender and Bussin,^32^ performance appraisals were confused with PM, which resulted in poor strategic focus. In Mboweni and Makhado,^33^ nurses only submitted appraisals for the entire quarter during the annual PMDS; there was no opportunity for feedback or continuous monitoring. The Gallup Group study concluded that knowing what is expected and the necessary tools to perform the job drives employee performance.^34^

Effective PMDS implementation tools depend on how seriously employees and managers use them.^35^ Therefore, managers/supervisors and employees must be trained,^35,36^ be proactive, familiar with the PMDS policy, and creatively think about PMDS’s effective implementation.

### Opportunities for collaboration and growth drive job satisfaction and achievement of the organisation’s goals

Building an employee engagement (EE)^32^ culture will leverage and drive performance.^33^ An engaged employee works positively to change behaviour and improve operational performance. Gruman and Saks highlighted the growing popularity of EE as the key to an organisation’s success.^37^ According to Robinson et al.^38^, EE is a positive attitude towards the organisation and its value. An engaged employee knows the business context and works with colleagues to improve job performance for the organisation’s benefit. The organisation must work to develop and nurture engagement, which requires a two-way relationship between employer and employee. An engaged employee is more than a nice thing; it has become a must.^38^ The MPST enabled employees’ engagement with the departmental strategic goals through inter-professional collaborations and learning, resulting in professional development and personal growth.

Schaufeli et al.^39^ explained that engaged employees can deal with demanding jobs and are connected to their work activities. In this study, although the participants experienced an initial sense of increased workload with the PMDS, its guided practice (MPST) added value, meaning, and purpose to work, triggering discomfort that resulted in professional and personal growth. This is unlike other studies^13,33,40^ in which employees developed negative feelings from favouritism, tension, competition amongst colleagues, a lack of meaning, and no opportunities to develop themselves. PMDS is perceived as punitive.^13^ The Public Service Commission toolkit 2007 for managing poor performance highlighted the primary orientation of PM as developmental.^12^ The MPST facilitated staff introspection, reflection, and identification of gaps, improved professional skills, innovation, collaboration with colleagues and other professionals, learning, and growth.

The unique experience of participants in this study was that working together among doctors and other healthcare professionals created a learning culture and improved the intra-facility referral system. The MPST provided guided self-directed learning and development, motivating participants. Robertson-Smith^41^ maintains that engaged employees feel empowered and grow in confidence in their ability to do their jobs. Collaboration among teams improves health outcomes, although it requires hard work. The lack of team effort in some studies^13, 33, 40^ was a significant barrier to PMDS implementation.

The MPST, an ongoing self-monitoring tool, allows employees to measure, identify gaps, and make decisions to correct and implement changes. This is cognisance with Menon’s^42^ descriptions, but unlike Lutwama et al.^43^ The Gallup Group identified critical employee expectations for engagement as employees wanting to know what is expected from them and the necessary tools to perform their jobs. They want to use their skills and talents and want recognition. It was vital that they feel valued. Feedback on their performance was necessary; they wanted learning opportunities and good relationships with their co-workers.^34^

Khan described that engaged employees express themselves physically, cognitively, and emotionally during role performances.^44^ Some participants expressed being more humane and empathetic, incorporating behaviour change that contributed to the patient’s positive care experience. Unlike this study, the lack of adequately prepared and skilled supervisors - managers merely complied - created a vacuum of meaning and purpose for the PMDS. Consequently, employees are not trained, PMDS is implemented with no goal-oriented strategies, and accountability is not enforced.^13^

### Authentic Leadership

Gruman and Saks^37^ linked the success and effectiveness of PM to the quality of leadership that engages, provides support, challenges, coaches, gives constructive feedback and includes employees in the organisation’s activities. As a clinical leader entrusted with governance, the SFP can aptly harness entrusted MOs’ abilities and broaden their knowledge and practice horizons through clear role identifications, support, and strategic tools. This is unlike the study by Govender and Bussin,^32^ where a lack of management and quality leadership negatively affected performance. Nxumalo et al. concluded that PMDS implementation at the local level depends on the ability of deployed managers to perform in a transforming and complex environment.^13^ This entails developing hard skills of planning, coordinating, monitoring, and soft skills in relationships and communication that promote collaborative and shared management.

The findings in this research regarding the meaning and purpose of work agree with Lockwood’s description of the benefits of a workplace culture that provides employees with a sense of meaning and purpose in their jobs.^27^ Work-role fit and support connect leaders and employees to shared beliefs and to attain organisational goals.

### Challenges

Bourne et al. described employees’ perception of the PM system as poor and likened it to the ‘big stick’ because they are not involved in the PA goal setting.^45^ This study resonates with what Madlabana et al.^25^ described regarding setting realistic goals with the staff at the coalface to ensure the standard of care is not compromised. Participants in this research stated their desire to be involved in setting achievable output targets that fit the context. This calls for self-introspection by managers.

Participants also accentuated other non-monetary rewards, such as certificates of appreciation. In the study by Govender and Bussin^32^, non-monetary recognition of the employees’ good work intrinsically motivates and boosts employees’ morale.^33^ Participants referred to ambiguity in the fairness of PMDS and monetary remuneration because they are aware of other units where employee scores do not equate to deserved performance, reflecting the manager’s lack of understanding of the PMDS. Also, some employees are driven by monetary rewards,^33,34^, and the PMDS lacks fairness, openness, and transparency and is subjective with unclear guidelines.^32,33^ It creates tension and competition amongst employees because rewards are not equally received, thus ruining relationships.^40^ Managers are fearful of giving unfavourable feedback and low scores.^13^

### Strength and limitations

This study provided broadened insights into the PMDS experiences of an unresearched cadre of healthcare professionals working in the PHC setting. An exploratory interpretive qualitative approach enabled participants to narrate and interpret their lived experiences. Introducing a monthly practice self-assessed performance and management tool could have influenced how this group of professionals experienced PMDS. Also, a stratified purposive sampling permitted the researcher to select all participants who lived the phenomenon studied and could provide rich, in-depth information. Seventeen MOs participated in this study, and saturation was attained because no new theme or information emerged with the fourth FGD. This study was done among MOs deployed to a particular region in the district. As a result, the findings may not represent other regions, districts, or low-middle-income countries. Further, the potential for social desirability bias may have been present because the researcher who conducted the FGDs was one of the two supervisors. To overcome this limitation, a research assistant was trained in the event of a power dynamic, which did not arise.

### Implications/recommendations

PMDS implementation tools, such as the MPST, with training and support for its use, were essential for participants to understand, value and make sense of the PMDS. A fit-for-purpose PMDS implementation tool will engage employees positively if used seriously by employees and managers/supervisors.

Professional and personal growth and job satisfaction were driven by being reflective clinicians, having a sense of meaning and purpose in work, and feeling that one contributes to the whole. Managers, who are leaders, are pivotal to leading clinical governance activities in the DHS. SFPs are tasked to lead and inspire managers and other clinicians in the district-based PHC health system to attain expected standards. Leadership and governance have been incorporated as learning outcomes for Family Physicians in training. The creation of posts and appointment of SFPs in the CHC and district hospitals will strengthen the DHS.

In the South African context of public-private disparity in the distribution of doctors, SFPs would play a significant role in attracting and retaining the right skills mix in doctors deployed to the PHC setting.

Participants propose management strategies, such as non-monetary awards or recognition of staff performance, that managers and supervisors should consider.

## Conclusion

This is the first study on doctors’ PMDS experiences in the researcher’s setting. The experiences of this cadre of healthcare professionals confirm some challenges faced by the South African PMDS. Moreover, participants’ experiences provided an in-depth and rich understanding of the how and benefits of PMDS implementation in a PHC setting with complex networks and relationships. The necessity of an implementation tool and clear leadership cannot be overlooked.

## Data Availability

Data cannot be shared publicly because it is the university's intellectual property. Data are available from the University of Pretoria Institutional Data Access / Ethics Committee (contact via)

## Acknowledments

The author acknowledges and appreciates the medical officers who participated in this study and colleagues on the peer-review team.

## References

1. Department of Public Service and Administration. SMS handbook, Chapter 4. Performance management and development [Internet]. c2006 [cited 2023 Apr 7]. Available from: https://www.dpsa.gov.za/dpsa2g/documents//sms/pmds/ch_4.pdf

2. World Health Organization [Internet]. The world health report: 2006: working together for health [Internet]. c2006 [cited 2023 Jun 1]. Available from: https://apps.who.int/iris/handle/10665/43432

3. World Health Organization [Internet]. Transforming and scaling up health professionals’ education and training. Guidelines. c2013 [cited 2023 Mar 16]. Available from: https://www.who.int/publications/i/item/transforming-and-scaling-up-health-professionals%E2%80%99-education-and-training

4. Public Service Commission. Evaluation of the effectiveness of the Performance Management and Development System for the Public Service [Internet]. c2018 [cited 2023 Jun 7]. Available from: https://www.psc.gov.za/documents/reports/2018/PMDS%20Report%20_%20Final.pdf

5. Naledi T, Barron P, Schneider H. Primary Health care in SA since 1994 and implications of the new vision for PHC re-engineering. South African Health Review [Internet]. 2011 [cited 2023 Apr 5];17-28. Available from: https://www.hst.org.za/publications/South%20African%20Health%20Reviews/2%20Primary%20Health%20Care%20in%20SA%20since%201994%20and%20%20%20Implications%20of%20the%20New%20Vision%20for%20PHC%20Re-engineering%20SAHR%202011.pdf

6. National Development Plan 2030. Our Future-make it work [Internet]. c2011 [cited 2023 Mar 23]. Available from: https://www.gov.za/sites/default/files/gcis_document/201409/devplan2.pdf

7. National Department of Health. A clinical governance handbook for District Clinical Specialist Teams [Internet]. c2014 [cited 2023 June 1]. Available from: https://knowledgehub.health.gov.za/system/files/elibdownloads/2023-04/Handbook%252520for%252520DCSTs%2525202014.pdf

8. Mash R, Von Pressentin KB. Strengthening the district health system through family physicians [Internet]. S Afr Health Rev [Internet]. 2018 series; 2018 [cited 5 Apr 2023];5(4):33-40. Available from: https://journals.co.za/doi/epdf/10.10520/EJC-1449142b2d

9. Public Service Commission. Chapter 7. Role of performance management [Internet]. [cited 2022 Dec 21]. Available from: https://www.psc.gov.za/conferences/dev-state-conference/Dev%20State%20Papers/Chapter%207%20Perf%20Man.pdf.

10. Department of Public Service and Administration. Determination and Directive on the Performance Management and Development System of employees other than members of the Senior Management Service for implementation with effect from 1 April 2018 [Internet]. [cited 2023 Apr 3]. Available from: https://www.psc.gov.za/conferences/dev-state-conference/Dev%20State%20Papers/Chapter%207%20Perf%20Man.pdf

11. Department of Public Service and Administration. GNR.877 of 29 July 2016: Public Service Regulation [Internet]. c2016 [cited 2023 Apr 3. Available from: https://www.dpsa.gov.za/dpsa2g/documents/acts&regulations/regulations2016/Public%20Service%20Regulation%202016%20as%20at%201%20November%202023.pdf

12. Republic of South Africa. Public Service Commission. Toolkit for the management of poor performance in the Public Service [Internet]. c2007 [cited 2024 Jan 18]. Available from: https://www.psc.gov.za/documents/docs/guidelines/K-6228%20PSC_Management%20of%20poor%20performance_Final.pdf

13. Nxumalo N, Goudge J, Gilson L, Eyles J. Performance management in times of change: experiences of implementing a performance assessment system in a district in South Africa. Int J Equity Health [Internet]. 2018 Sept [cited 2024 Jan 18];17(141):1-14. Available from: 10.1186/s12939-018-0857-2.

14. Grossoehme DH. Research methodology overview of qualitative research. J Health Care Chaplain [Internet]. 2014 Oct [cited 2024 Feb 1];20(3): 109-22. Available from: https://pmc.ncbi.nlm.nih.gov/articles/PMC4609437/pdf/nihms-726694.pdf.

15. Renjith V, Yesodharan R, Noronha JA, Ladd E, George A. Qualitative methods in Health care research. Int J Prev Med [Internet]. 2021 Feb [cited 2023 Jan 24]:12: 20. Available from: https://pmc.ncbi.nlm.nih.gov/articles/PMC8106287/pdf/IJPVM-12-20.pdf

16. Merriam SB. Introduction to qualitative research. In Merriam, Sharon and Associates, eidtors. Qualitative Research in Practice.. San Francisco: Jossey-Bass; 2002.

17. National Department of Health. Integrated clinical services management [Internet]. c2015 [cited 2024 Aug 3]. Available from: https://knowledgehub.health.gov.za/system/files/elibdownloads/2023-04/Integrated%252520Clinical%252520Services%252520Management%252520%252520Manual%2525205th%252520June%252520FINAL.pdf

18. Bradshaw C, Atkinson S, Doody O. Employing a qualitative description approach in health care research. Glob Qual Nurs Res [Internet]. 2017 Nov [cited 2023 Jan 24]; 4:1–8. Available from: https://pmc.ncbi.nlm.nih.gov/articles/PMC5703087/19.

19. Burdine JT, Thorne S, Sandhu G. Interpretive description: A flexible qualitative methodology for medical education research [Internet]. Med Educ [Internet]. 2021 Mar [cited 2023 Jan 24];55(1):336–343. Available from: https://asmepublications.onlinelibrary.wiley.com/doi/10.1111/medu.14380. Subscription required.

20. Cypress B. Rigor, reliability and validity in qualitative research: perspectives, strategies, re-conceptualization and recommendations. Dimens Crit Care Nurs [Internet]. 2017 Jul/Aug [cited 2024 Jan 30];36(4):253-63. Available from: file:///C:/Users/nixsm/Downloads/rigor_or_reliability_and_validity_in_qualitative.6.pdf

21. Lincoln YS, Guba EG. Naturalistic inquiry [Internet]. California: Sage; 1985

22. Stahl NA, King JR. Expanding approaches for research: understanding and using trustworthiness in qualitative research. J Dev Educ [Internet]. 2020 [cited 2024 Jan 30]; 44(1):26-8. Available from: https://files.eric.ed.gov/fulltext/EJ1320570.pdf

23. Moser A, Korstjen I. Series: practical guidance to qualitative research. Part 1: Introduction. Eur J Gen Pract [Internet]. 2017 Nov[cited 2023 Sep 5];23(1):271–3. Available from: https://www.tandfonline.com/doi/epdf/10.1080/13814788.2017.1375093?needAccess=true

24. Aguinis H. Performance management. 3^rd^ ed. New Jersey: Pearson; 2013

25. Madlabana CZ, Petersen I. Performance management in primary healthcare: nurses’ experiences. Curationis [Internet]. 2020 Apr [cited 2024 Jan 18]; 43(1):1–11. Available from: https://curationis.org.za/index.php/curationis/article/view/2017/2724

26. Mashego RH, Skaal L. Knowledge and practices of supervisors on the performance management and development system at rural primary health care facilities in Limpopo Province. Afr J Prim Health Care Fam Med [Internet]. 2016 Dec [cited 2024 Jan 18]; 8(1):1–5. Available from: https://pmc.ncbi.nlm.nih.gov/articles/PMC5153408/pdf/PHCFM-8-1236.pdf

27. Lockwood NR. Leveraging employee engagement for competitive advantage: HR’s strategic role. SocHum Res Manag Res Quart [Internet]. 2007 Mar [cited 2024 Jan 18];52(3):1–11. Available from: https://www.proquest.com/docview/205067594?sourcetype=Trade%20Journalsz.

28. Sambo NM, van der Walt G. A strategic framework for public performance management and development system implementation. Admin Pub [Internet]. 2022 Jun [cited 2024 Jan 18]30(2):86–106. Available from: https://journals.co.za/doi/epdf/10.10520/ejc-adminpub_v30_n2_a8

29. Melnyk SA, Bititci U, Platts K, Tobias J, Anderson. Is Performance Measurement and Management Fit for the Future? Manag Account Res [Internet]. 2014 Jun[cited 2024 Jan 30]; 25(2):173–86. Available from: https://www.sciencedirect.com/science/article/abs/pii/S1044500513000723 Subscription Required.

30. Smither JW, London M. Performance management: putting research into action. The Professional Practice series. John Wiley & Sons;2009.

31. DeNisi AS, Pritchard RD. Performance appraisal, performance management and improving individual performance: a motivational framework. Manag Organ Rev [Internet]. 2006 [cited 2024 Jan 30];2(2):253–277. Available from: https://econpapers.repec.org/article/blamgorev/v_3a2_3ay_3a2006_3ai_3a2_3ap_3a253-277.htm Subscription Required.

32. Govender M, Bussin MHR. Performance management and employee engagement: a South African perspective [Internet]. SA J Hum Resour Manag. 2020 Jun [cited 2024 Jan 13];18:1–19. Available from: https://sajhrm.co.za/index.php/sajhrm/article/view/1215/2067

33. Mboweni SH, Makhado L. Professional nurses’ lived experiences regarding the performance management system in the Mopani district. Curationis [Internet]. 2017 Oct [cited 2024 Jan 19];40(1):1–9. Available from: https://pmc.ncbi.nlm.nih.gov/articles/PMC6091791/pdf/CUR-40-1631.pdf

34. Gopal A. Worker disengagement continues to cost Singapore. Gallup Manag J [Internet]. 2006 May [cited 2024 Jan 19]. Available from: https://static1.squarespace.com/static/578c4a19197aea41c7eed064/t/59af9d06a9db09ad7b9c3e0b/1504681224814/worker+disengagement+continues+to+cost+singapore+(gallup+management+journal).pdf

35. Pulakos ED. Performance management: a roadmap for developing, implementing and evaluating performance management systems. Society for Human Resource Management [Internet]. c2004 [cited 2024 Feb 8]. Available from: https://www.studocu.com/en-za/document/regent-business-school/human-resources-management-and-labour-relation/performance-management-book-idm/26696449

36. Malefane SR. Is it really performance that is measured? A reflection of the South African Government’s performance management system. J US-China PubAdmin [Internet]. 2010 Aug[cited 2024 Feb 1];7(8);1–12. Available: https://uir.unisa.ac.za/bitstream/handle/10500/23764/Is%20it%20really%20performance%20that%20is%20measured%20A%20reflection%20of%20the%20South%20African%20governments%20performance%20management%20system.pdf?sequence=1

37. Gruman JA, Saks AM. Performance management and employee engagement. Hum Res Manag Rev [Internet]. 2011 Jun [cited 2024 Feb 1];21(2):123–36. Available from: https://www.sciencedirect.com/science/article/abs/pii/S1053482210000409 Subscription Required.

38. Robinson D, Perryman S, Hayday S. The drivers of employee engagement. Institute for Employment Studies [Internet]. c2004 [cited 2024 Feb 1]. Available from: https://instituteforpr.org/employee-engagement-5/#:~:text=A%20sense%20of%20feeling%20valued%20and%20involved%20was,organization%20was%20concerned%20for%20their%20health%20and%20well-being.

39. Schaufeli WB, Salanova M, González-Romá V, Bakker AB. The measurement of engagement and burnout: a two sample confirmatory factor analytic approach. Journal of Happiness Studies [Internet]. 2002 Mar [cited 2024 Feb 1];3(1):71–92. Available from: https://www.scribd.com/document/223058110/2002-The-Measurement-of-Burnout-and-Engagement-a-Confirmatory-Factor-Analytic-Approach

40. Tyokwe B. Naicker V. The effectiveness of a performance system at a South African public hospital in Cape Town. African Pub Serv DelPer Rev [Internet]. 2021 May [cited 2024 Jan 18];8(1):1–10. Available from: https://www.academia.edu/95431165/The_effectiveness_of_a_performance_management_system_at_a_South_African_public_hospital_in_Cape_Town

41. Robertson-Smith G, Markwick C. Employee engagement: a review of current thinking. Institute for Employment Studies [Internet]. c2009 [cited 2024 Feb 8]. Available from: https://www.employment-studies.co.uk/resource/employee-engagement-review-current-thinking

42. Menon S. Employee empowerment: an integrative psychological approach. Appl Psychol [Internet]. 2001 [cited 2024 Feb 1]; 50(1):153–180. Available from: https://www.bwgriffin.com/gsu/courses/edur9131/activities/Menon_ST_2001_employee_empowerment_Applied_Psychology.pdf

43. Lutwama G, Roos J, Dolamo BL. Assessing the implementation of performance management of health care workers in Uganda. BMC Health Serv Res [Internet]. 2013 Sep [cited 2024 Feb 1]; 13(1): 1–12. Available from: https://bmchealthservres.biomedcentral.com/articles/10.1186/1472-6963-13-355

44. Kahn WA. Psychological conditions of personal engagement and disengagement at work. Acad Manage J [Internet]. 1990 Nov [cited 2024 Feb 8]; 33(4): 692–724. Available from: https://www.jstor.org/stable/256287

45. Bourne M, Neely A, Platts K, Mills J. The success and failure of performance measurement initiatives: perceptions of participating managers. Int J Oper Prod Manag [Internet]. 2002 Nov [cited 2024 Feb 1]; 22(1): 1288–1310. Available from: https://www.emerald.com/insight/content/doi/10.1108/01443570210450329/full/html Subscription Required.

